# Estimation of the percentages of asymptomatic patients and undiagnosed patients of the novel coronavirus (SARS-CoV-2) infection in Hokkaido, Japan by using birth-death process with recursive full tracing

**DOI:** 10.1101/2020.04.06.20053934

**Authors:** Takuma Tanaka, Takayuki Yamaguchi, Yohei Sakamoto

## Abstract

Estimating the percentages of undiagnosed and asymptomatic patients is essential for controlling the outbreak of SARS-CoV-2, and for assessing any strategy for controlling the disease. In this paper, we propose a novel analysis based on the birth-death process with recursive full tracing. We estimated the numbers of undiagnosed symptomatic patients and total infected individuals per diagnosed patient before and after the declaration of the state of emergency in Hokkaido, Japan. The median of the estimated number of undiagnosed symptomatic patients per diagnosed patient decreased from 1.9 to 0.77 after the declaration, and the median of the estimated number of total infected individuals per diagnosed patient decreased from 4.7 to 2.4. We will discuss the limitations and possible expansions of the model.

## Introduction

The novel coronavirus (SARS-CoV-2) spread to the most populated areas of the world in the first few months of 2020. In Japan, the first case was reported on January 16th, 2020; on March 31st, the number of cases increased to 2122 [1]. In Hokkaido, the largest prefecture of Japan, the first case was reported on January 28th, 2020 (Fig 1). The Hokkaido government declared a state of emergency on February 28th and lifted it on March 19th. The state of emergency was not legally binding, and the government asked the residents to stay home on the weekends. Until the state of emergency was lifted, a total of 157 cases were reported. Although the number of diagnosed cases is declining towards the end of March, the situation remains uncertain.

**Fig 1.**
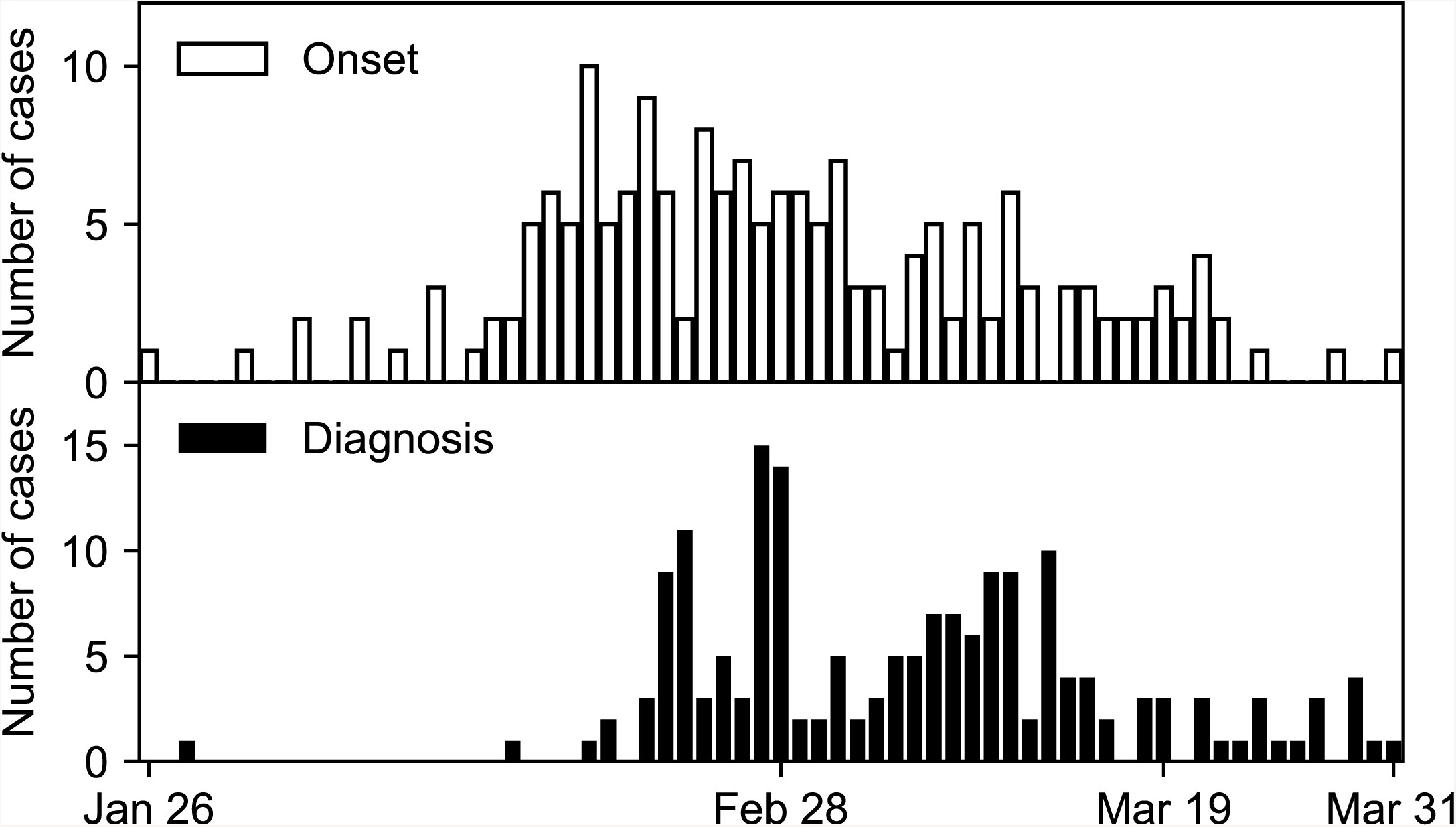
Epidemic curves of COVID-19 in Hokkaido, early 2020. The histograms of the date of onset (above) and the date of diagnosis (below) are shown.

To effectively control the spread of the infection, we need to know several parameter values characterizing the infection, such as the basic reproduction number, *R*_0_, the percentage of asymptomatic patients, and the fatality rate. One of the factors that complicate the decision making in disease control is the uncertainty in the percentage of asymptomatic patients. Several lines of evidence indicate that the virus can be transmitted by asymptomatic patients [2, 3], who may have facilitated the spread of SARS-CoV-2. To make the disease control strategy successful and less harmful to the society, the transmissibility of asymptomatic patients and the percentage of asymptomatic patients should be measured. Another complicating factor is the uncertainty in the percentages of diagnosed and undiagnosed patients. Most individuals infected with SARS-CoV-2 suffer from mild cold-like symptoms and recover without any medical intervention. They remain undiagnosed, disturbing how we monitor the number of total infected individuals. Thus, estimating the percentages of asymptomatic and undiagnosed patients is a challenging problem to be answered in epidemiology.

It would be useful if these values could be estimated by the information available in the early phase of the outbreak. Contact tracing is considered to be one of the effective measures, and health officials have conducted contact tracing of infected patients to prevent the spread of the virus infection for outbreaks of new or reemerging infections [4–9]. If an infected patient is found, health officials try to find other infected people among those who have come into contact with the infected patient. If any other infected patients are found, officials will repeat tracing from the newly found patients; and if not, tracing is stopped. Thus, a “cluster” of infected patients connected by the route of infection is constructed through the process of contact tracing. Simultaneously, the dates of onset and diagnosis are obtained for each diagnosed patient. As a result, contact tracing can provide detailed qualitative and quantitative information about diagnosed patients from the infectious disease transmission network of diagnosed and undiagnosed patients. Analyzing this network with an appropriate model may enable us to estimate the parameter values of the infection.

One promising model for contact tracing is a stochastic model based on the birth-death process, which is a formulation of branching processes [10–17], because the number of cases in the early phase stochastically fluctuates and the widely used differential-equation-based models are inapplicable. In birth-death processes, a sequence of infectious events generates a tree whose nodes are infected patients and edges are infection routes (Fig 2). When a patient recovers (Fig 2, dotted circle), the corresponding node and its edges are removed from the tree, which is split into two. The infectious disease transmission network is composed of trees. A connected component of the network is referred to as “cluster” in this paper. Contact tracing corresponds to finding a node in the tree and removing nodes connected to the first node found (Fig 2, gray filled circle). There are various types of contact tracing based on how to choose nodes to be removed, including backward, forward, or full tracing, and recursive or one-step tracing [17]. We consider only recursive full tracing, that is, all nodes directly and indirectly connected to the first found node are removed (Fig 2, dashed rounded rectangle). We propose an analysis based on the birth-death process with recursive full tracing that takes advantage of information obtained by contact tracing to estimate epidemiological parameter values with a small set of data. We focus on the contact tracing of infected individuals in Hokkaido. The present analysis uses the distributions of the cluster size and patients’ time from onset to diagnosis, which are released by the health officials, to estimate the model parameters. Our approach directly models the stochastic dynamics, which is an inherent property of the early phase of the outbreak.

**Fig 2.**
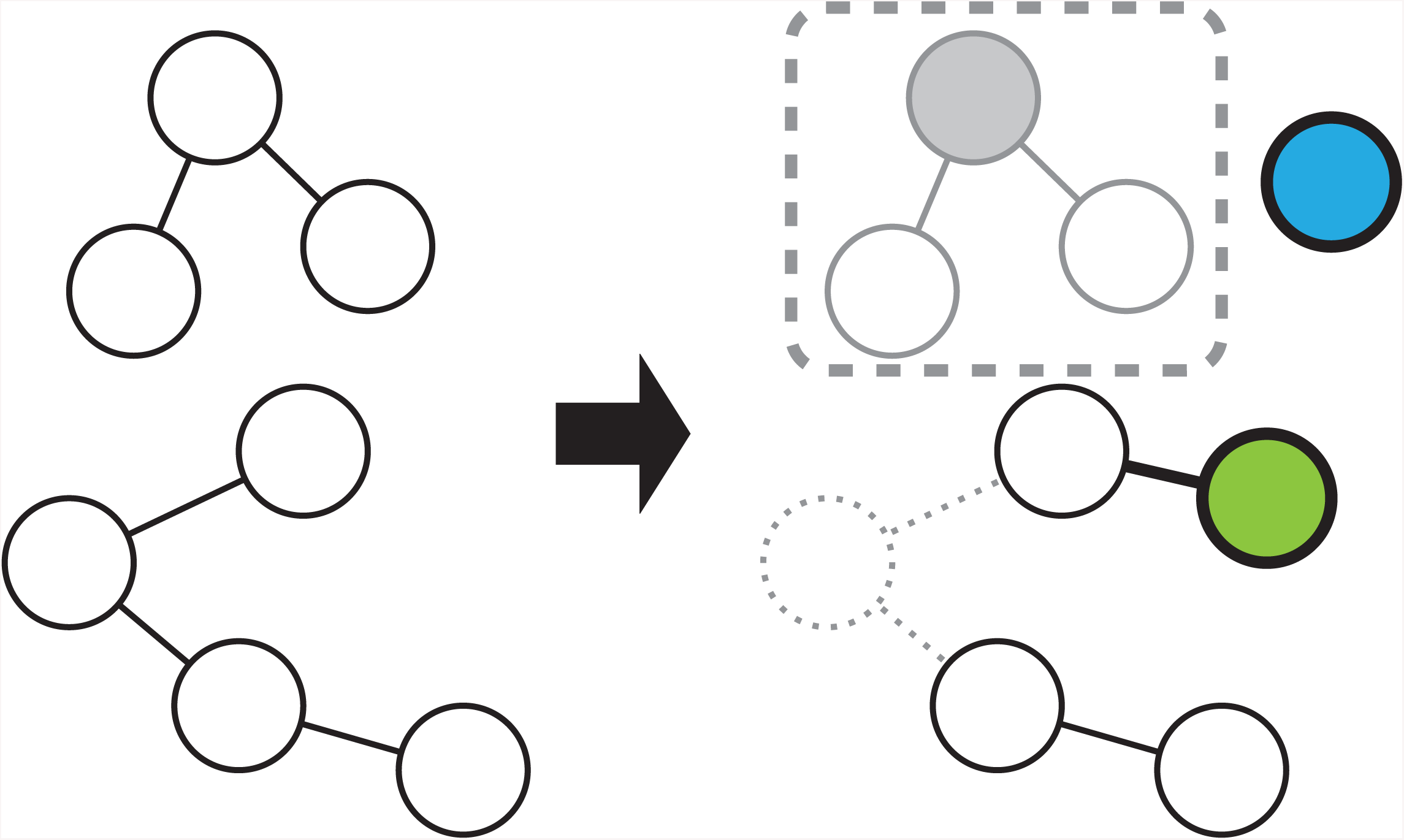
Infection, recovery, and diagnosis in the birth-death process with recursive full tracing. A network with two clusters is shown on the left side, and the network after the progression of events is shown on the right side. The nodes represent symptomatic patients, and two nodes are connected by an edge if one has infected the other. The nodes can recover and be removed from the network (dotted circle/lines), or infect and connect to a new node (green circle). A new node without edges can be generated in the network (blue circle). A node can be diagnosed (gray filled circle), and the nodes in the same connected component (gray open circles) are removed from the network and counted among diagnosed symptomatic clusters (dashed rounded rectangle). Nodes and edges removed from the network are indicated in gray, and those newly generated in the network are indicated by bold lines.

This paper is organized as follows. In the Methods section, we summarize the SARS-CoV-2 infection in Hokkaido and divide it into those before and after the declaration of the state of emergency. We classify patients into symptomatic and asymptomatic, and diagnosed and undiagnosed. We explain what corresponds to diagnosed and undiagnosed patients in the present model. We describe the formulation of the model and the details of the simulations. In the Results section, we estimate the parameter values and the number of asymptomatic and undiagnosed patients. In the Discussion section, we relate our results with previous studies and discuss the limitations and possible expansions of the model.

## Methods

This paper reports the analysis of the SARS-CoV-2 infection in Hokkaido, Japan [18]. In Hokkaido, all cases had not traveled abroad recently except for three cases, which include a tourist from Wuhan, China. We concentrate our analysis on the cases whose onset was prior to the lifting of the state of emergency. We excluded the cases with an unknown onset and asymptomatic patients from the analysis. If the date of the diagnosis of a case was not reported, it was assumed to coincide with the date of the announcement. S1 Table summarizes the case reports released by the Novel Coronavirus Response Headquarters of the Hokkaido government until April 2nd, 2020.

We represent the patients with nodes and their contacts with edges in a network. If two patients were in close contact with each other, the corresponding nodes are connected by an edge. The network consists of distinct connected components, which we refer to as clusters. Sporadic patients are regarded as size-1 clusters. Fig 3 shows the clusters with sizes larger than 2 in Hokkaido. There are 76 size-1 clusters and 20 size-2 clusters along with the clusters shown in Fig 3 (S1 Table).

**Fig 3.**
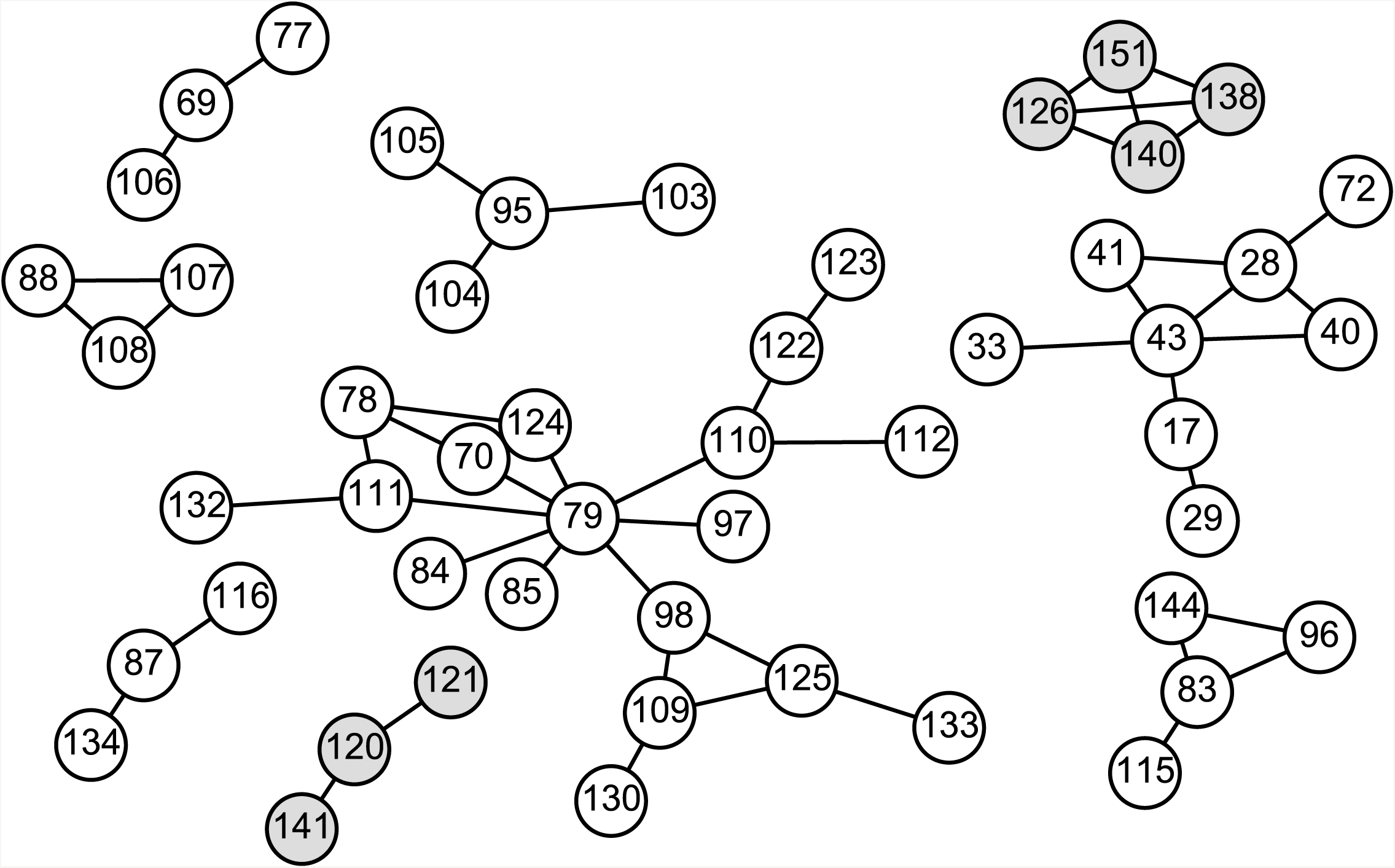
Contact network between patients in Hokkaido. The white and gray circles represent the patients in dataset 1 and 2, respectively. The numbers in the circles are the case IDs. Two circles are connected by an edge if these two patients were in close contact with each other.

All cases were divided into datasets 1 and 2 according to which cluster they belong to. If the earliest onset of the cases in a cluster was prior to the declaration of the state of emergency, this cluster was included in dataset 1; if the earliest onset was between the declaration and the lifting thereof, it was included in dataset 2. Datasets 1 and 2 contain 78 and 30 clusters, respectively. Because the declaration of the state of emergency might have changed the behavior of residents and health officials in Hokkaido, we compared the data before the declaration, dataset 1, and the data between the declaration and the lifting thereof, dataset 2.

The patients included in datasets 1 and 2 were all diagnosed and mostly symptomatic. However, not all of the individuals infected with SARS-CoV-2 were diagnosed and symptomatic; they can be classified into diagnosed symptomatic, diagnosed asymptomatic, undiagnosed symptomatic, and undiagnosed asymptomatic groups. The diagnosed symptomatic group consists of those who developed symptoms and were diagnosed, or those who were found in contact tracing. All individuals belonging to this group were covered by our datasets. Although the diagnosed asymptomatic group was also included in the datasets, we ignored this group because this group included only two individuals. The undiagnosed symptomatic group is comprised of individuals who were infected and developed symptoms, but recovered or died without being diagnosed. This group is not directly observable, and thus its percentage is one of the parameters we tried to estimate with the model, which takes this group into account. The undiagnosed asymptomatic group is not directly observable either. It has been suggested that a percentage of SARS-CoV-2 carriers do not develop symptoms but can infect others [2, 19]. We did not explicitly incorporate the asymptomatic group into the model but estimated its percentage.

We modeled the spread of SARS-CoV-2 with a variant of the continuous-time birth-death process, referred to as the birth-death process with recursive full tracing [17]. Birth-death processes have been used to model infectious diseases and population dynamics [10, 11, 16, 17]. Continuous-time birth-death processes consist of a network whose nodes and edges are continually generated and removed (Fig 2). The lifetime of a node is a random variable drawn from the exponential distribution with the scale parameter 1*/γ*. During its lifetime, a node gives birth to nodes according to a Poisson point process with the stationary rate *β*′ and is connected to these offspring nodes (Fig 2, green circle). When the lifetime of a node ends, the node and its edges are removed from the network (Fig 2, dotted circle). For the infection dynamics of SARS-CoV-2, the nodes and their birth and death can be regarded as symptomatic patients and their onset of symptoms or infection and recovery from the disease. In the limit of an infinite number of nodes, the dynamics of the number of nodes of birth-death processes are approximated by the SIR model. Asymptomatic infected individuals are not included in this model, and the incubation period is ignored.

The birth-death process with recursive full tracing incorporates the diagnosis and quarantine of patients in addition to the features of continuous-time birth-death processes. The time from infection to diagnosis of a node is a random variable drawn from the exponential distribution with the scale parameter 1*/κ*. If the diagnosis occurs earlier than the recovery, the node is removed from the network at the diagnosis, representing the quarantine of the diagnosed patient (Fig 2, gray filled circle). At the same time, the nodes in the connected component containing the diagnosed node are also removed from the network, which corresponds to the contact tracing of the infected individuals (Fig 2, gray open circles). Infections that are to be caused by these removed nodes later than the removal are abolished because the diagnosed individuals are quarantined. The connected component of the diagnosed individuals corresponds to the clusters in the datasets (Fig 2, dashed rounded rectangle). The nodes in the connected component are counted among diagnosed symptomatic groups. The recovery of a node disconnects its neighboring nodes. For example, if the green node in Fig 2 is diagnosed, the cluster of size 2 but not of size 4 is reported. A node is not diagnosed if it has been already removed. If a node recovers before it is diagnosed, this node is counted among undiagnosed symptomatic groups.

The simulation of the model is implemented as follows. Nodes without edges are generated in the network according to a Poisson point process with the stationary rate *λ* = 10^−5^ (Fig 2, blue circle). The value of *λ* is inconsequential if it is small enough to allow for observing the cluster size and time from onset to diagnosis distribution at the steady state. On the generation of node *i* at time 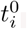, its time of recovery 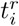 and time of diagnosis 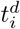 are assigned as

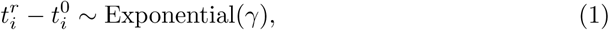

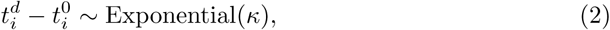

where the recovery rate *γ* and the diagnosis rate *κ* are positive constants. During 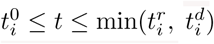, node *i* generates new nodes and connects to them according to a Poission point process with the stationary rate *β*′ > 0. At 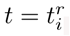, if node *i* is present in the network, node *i* and its edges are removed from the network. At 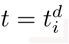, if node *i* is present in the network, the nodes in the same connected component containing node *i* and their edges are removed from the network.

Let us note that *β*′ is the rate of infection giving rise to symptomatic patients, not the rate of infection giving rise to symptomatic and asymptomatic patients. This is because all nodes in the model are capable of being diagnosed, which is not the case with asymptomatic individuals. Hence, *β*, the rate of infection giving rise to any type of patient is greater than *β*′. For *κ* = 0, the probability that a node directly infects *n* nodes, that is, the probability that a symptomatic patient directly infects *n* symptomatic patients, follows

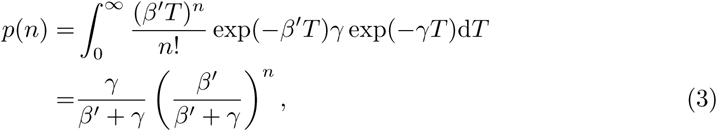

whose expected value is *β*′*/γ*. Similarly, the basic reproduction number *R*_0_ is given by *β/γ*. Thus, *β* must be greater than *γ* because the number of reported cases is steadily increasing, that is,

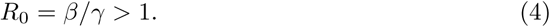

Throughout the simulations reported in this paper, *γ* was fixed to 1*/*14 [20–22].

We performed the approximate Bayesian computation of the posterior distribution of *κ* and *β*′ given the average cluster size and the average time from onset to diagnosis. We drew *β*′ from U(0.001, 0.2) and *κ* from U(0.001, 0.12) and accepted the parameter sets with which the average cluster size was identical to 126/78 for dataset 1 and 43/30 for dataset 2, and the average time from onset to diagnosis lied within ±1 days of 9.3 for dataset 1 and 6.6 for dataset 2. The cluster size is defined as the number of nodes of the cluster. The average time from onset to diagnosis is defined as the average of 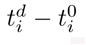 where *i* runs over the nodes that are diagnosed first in the cluster.

In each run of the simulation, we removed *C*_0_ + *C* clusters of diagnosed symptomatic patients, of which the first *C*_0_ = 100 clusters were discarded to eliminate the dependence of the results on the initial condition and used the following *C* = 78 (dataset 1) and *C* = 30 (dataset 2) clusters as the simulated clusters of patients in Hokkaido. This procedure is justified by the fact that the average cluster size of birth-death processes converges to its steady-state value on a timescale of 1*/β* and 1*/γ* [11]. This fact also suggests that the properties of clusters in the early phase of the spreading of SARS-CoV-2 can be described by the steady-state of the model.

The ratio of undiagnosed symptomatic patients to diagnosed symptomatic patients can be estimated by the number of nodes that recover without being diagnosed in a period divided by the number of diagnosed nodes that recover in the same period. We used the period between the removal of the *C*_0_-th cluster and the removal of the *C*_0_ + *C*-th cluster to calculate the ratio. This period is referred to as the target period in the following. The ratio of the number of symptomatic and asymptomatic patients to the number of symptomatic patients is *β/β*′. Because *β* > *γ*, which follows from Eq 4, and *β* > *β*′, the lower bound of the number of all infected individuals that recover in the target period is estimated by the number of diagnosed and undiagnosed symptomatic patients that recover in the period multiplied by max(*γ, β*′)*/β*′. The estimates presented in this paper are rounded to two significant digits.

## Results

We performed simulations with randomly generated 100 000 parameter sets and accepted the parameter sets that replicated the average cluster size and the average time from onset to diagnosis. We chose these two indices as those that are used to fit the model parameters for two reasons. First, these can be obtained without using sophisticated techniques. Second, these two indices enable us to determine the parameter values of *β*′ and *κ*. Before applying the parameter estimation to datasets 1 and 2, we tested it for the indices obtained from a simulation run with *β*′ = 0.1 and *κ* = 0.05. The orange crosses, blue triangles, and filled circles in Fig 4A indicate the parameter sets that replicated the average cluster size, the average time from onset to diagnosis, and both together, respectively. This figure shows that the filled circles are concentrated on the intersection of the bands of crosses and triangles. The 95% credible intervals for *β*′ and *κ* were [0.081, 0.14] and [0.033, 0.081], respectively, which successfully contain the parameter set that was used in the original simulation.

**Fig 4.**
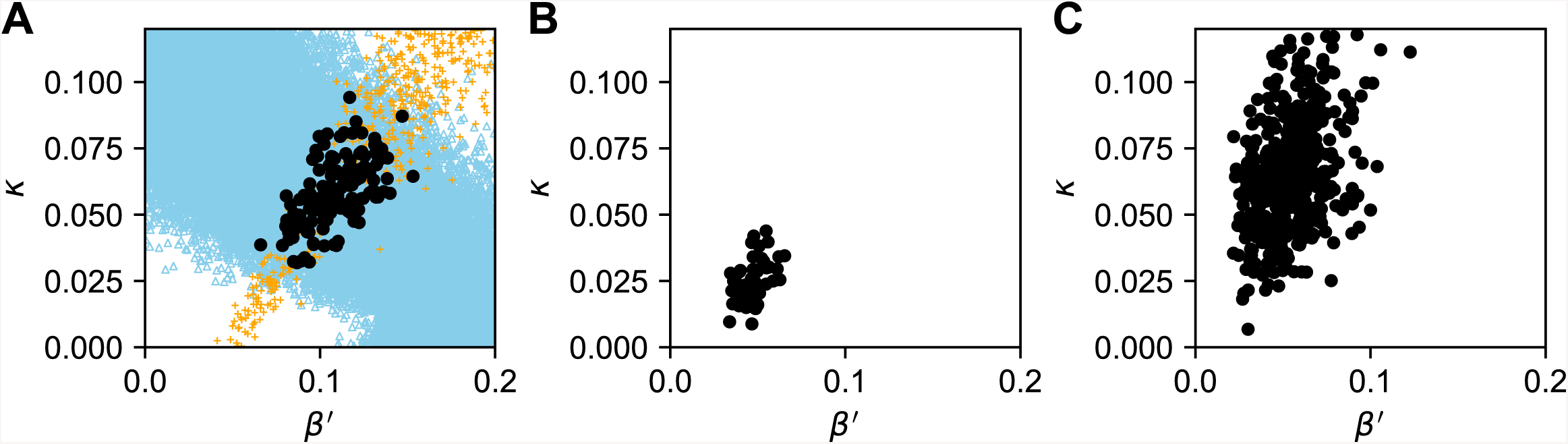
The estimated parameter sets for the simulated data and datasets 1 and 2. The parameter sets that replicated both of the average cluster size and the average time from onset to diagnosis of a simulation run with *β*′ = 0.1 and *κ* = 0.05 (A), dataset 1 (B), and dataset 2 (C) are indicated by the filled circles. Panel A also shows the parameter sets that replicated the average cluster size (orange crosses) and the average time from onset to diagnosis (blue triangles).

Figs 4B and 4C present the parameter sets that replicated the average cluster size and the average time from onset to diagnosis of datasets 1 and 2, respectively. The 95% credible intervals for *β*′ and *κ* were [0.035, 0.062] and [0.012, 0.041] for dataset 1, and [0.027, 0.092] and [0.028, 0.11] for dataset 2. In dataset 1, the median of the estimated value of *κ*, 0.024, was far less than *γ*, suggesting that most of the symptomatic patients were not diagnosed before their recovery. The median of the estimated value of *κ* in dataset 2, 0.063, implies that a larger percentage of symptomatic patients were diagnosed in dataset 2.

To examine the number of undiagnosed symptomatic patients, we calculated the number of nodes that recovered without being diagnosed in the target period. Fig 5A and C show the number of undiagnosed symptomatic patients per diagnosed symptomatic patient for dataset 1 and 2, respectively. The 95% credible intervals were [1.0, 3.6] (median 1.9) for dataset 1 and [0.34, 2.0] (median 0.77) for dataset 2. The lower bound of the number of all infected individuals is estimated by the number of diagnosed and undiagnosed symptomatic patients multiplied by max(*γ, β*′)*/β*′ (Fig 5B, D). The 95% credible intervals of the total number of infected individuals per diagnosed patient were [2.6, 8.6] (median 3.7) for dataset 1 and [1.4, 5.8] (median 2.4) for dataset 2, the former of which is consistent with a previous estimate, 1*/*0.14 [23]. These estimates suggest that around half of infected individuals remain asymptomatic, which is consistent with a previous estimate [19]. The 95% credible intervals of the total numbers of infected individuals who recovered before the declaration of the state of emergency and those who recover between the declaration and lifting were [330, 1100] (median 600) and [61, 250] (median 100), respectively.

**Fig 5.**
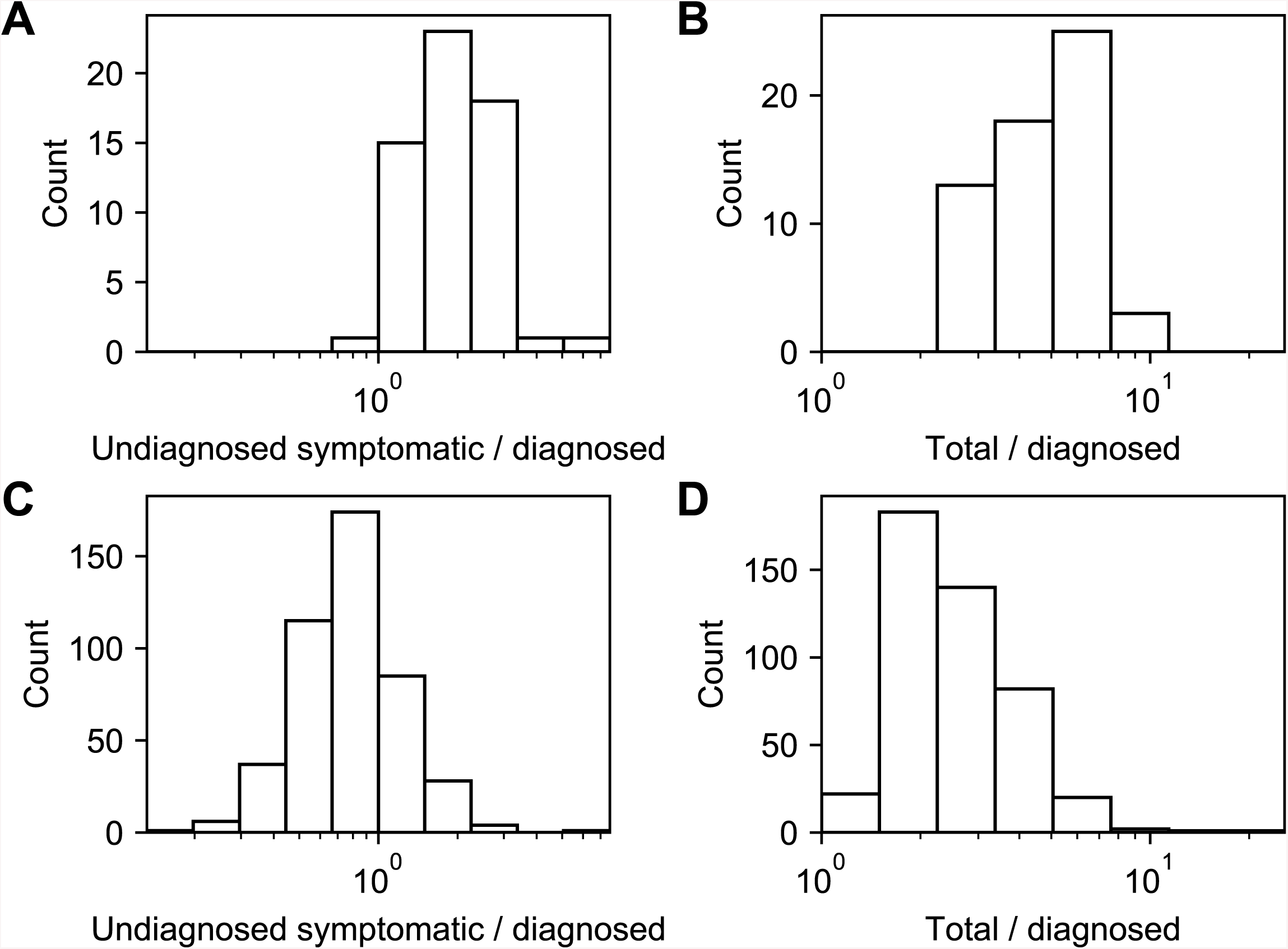
The estimated numbers of undiagnosed symptomatic patients (A, C) and asymptomatic patients (B, D) per diagnosed symptomatic patient. Panels A and B are for dataset 1, and panels C and D are for dataset 2.

## Discussion

In this paper, we have formulated a model to describe the spreading of infection and the quarantine of infected individuals, and estimated the number of undiagnosed symptomatic and asymptomatic COVID-19 patients in Hokkaido. The estimated percentages of undiagnosed symptomatic and asymptomatic patients coincided with previous studies [23, 24]. The estimated total number of patients that recovered before the declaration of a state of emergency was also consistent with a previous study, which estimated that there were 940 patients in Hokkaido on February 25th [25]. One of the previous studies approximated the time evolution of the number of infected individuals with differential equations [23], while another estimated the number of asymptomatic patients by using RT-PCR (reverse transcription polymerase chain reaction) test results of evacuees from Wuhan, China on chartered flights [24]. The present analysis focuses on the stochastic dynamics of a discrete number of infected individuals. Thus, the size distribution of clusters, which is a piece of information available in the early phase of the pandemic but difficult to use in differential-equation-based models, can be utilized by the model. Although the methods of the previous reports and ours are completely different, quantitative agreement between them suggests the effectiveness of these approaches.

There are several reasons we have chosen the cases in Hokkaido as the subject of this paper. Hokkaido is an island isolated from the other regions of Japan. In other words, we can assume that a relatively small percentage of the population commutes between Hokkaido and other parts of the world. This makes Hokkaido an ideal subject of the investigation. Until March 20th, one day after the lifting of the state of emergency, 1549 out of 1707 individuals tested with RT-PCR turned out to be negative for SARS-CoV-2 [26], indicating that extensive contact tracing was performed. On the other hand, among the 158 cases diagnosed until March 19th in Hokkaido, only two were asymptomatic. This suggests that in most contact tracing in Hokkaido, RT-PCR tests for SARS-CoV-2 were conducted only on symptomatic patients because of restricted resources. The claim by the local government that test capability was strengthened after the declaration of the state of emergency [27] is supported by a larger value of *κ* in dataset 2 than in dataset 1.

One of the features of the present model is its simplicity. The model has only three essential parameters. The simplicity of the model allowed us to estimate the number of asymptomatic and undiagnosed patients without using a large number of parameter values estimated by previous studies. In the early phase of the spreading of infectious diseases, this simple model can enable the estimation of the asymptomatic and undiagnosed patients despite limited data. Our approach can estimate the number of patients without using costly and time-consuming techniques such as RT-PCR.

Also, its simplicity might allow for an analytical solution. The model is an extension of the birth-death processes, which has been studied intensively. The birth-death processes with contact tracing is analytically tractable [10, 11, 16, 17]. Because the model is simple enough, we may be able to obtain the analytical solution of the cluster size distribution and the expected time from onset to diagnosis. The analytical solution would enable us to efficiently estimate the parameter values.

The simplicity of the present model allows expansions in several ways. First, we assumed that *β*′ is a fixed value in this paper. This is justified by Fig 3, which shows that the largest number of individuals infected by an individual, that is, the largest degree of nodes, is eight, which is rather small. However, *β*′ may be heterogeneous. Heterogeneity in *β*′, which has been suggested for the coronavirus genus [28, 29], may explain the superspreader phenomenon. Also, the spread of SARS-CoV-2 might be more accurately modeled with the contact process on scale-free networks [30, 31]. The value of *β*′ might depend on the severity of the patient [23]. Second, the recovery rate *γ* may depend on the severity of the patient. Heterogeneity in *γ* can affect the estimated number of total infected individuals. Third, the incubation period, which is ignored in the present paper, might affect the size and structure of clusters [32]. Infectiousness in the incubation period should be included in the model [32]. Fourth, the stage of symptoms should be introduced into the model. Fig 6 suggests that the time from onset to diagnosis obeys a unimodal distribution with a peak at around 10 days, although the peak must be at 0 in the present model. Assuming that a mildly infected state stochastically develops into a severely infected state would explain this time course. Fifth, recursive full tracing might be unrealistic because some of the symptomatic patients can be missed in contact tracing. Introducing stochasticity into contact tracing can enable a more precise modeling of clusters. These extensions would be useful in monitoring and controlling the spread of SARS-CoV-2.

**Fig 6.**
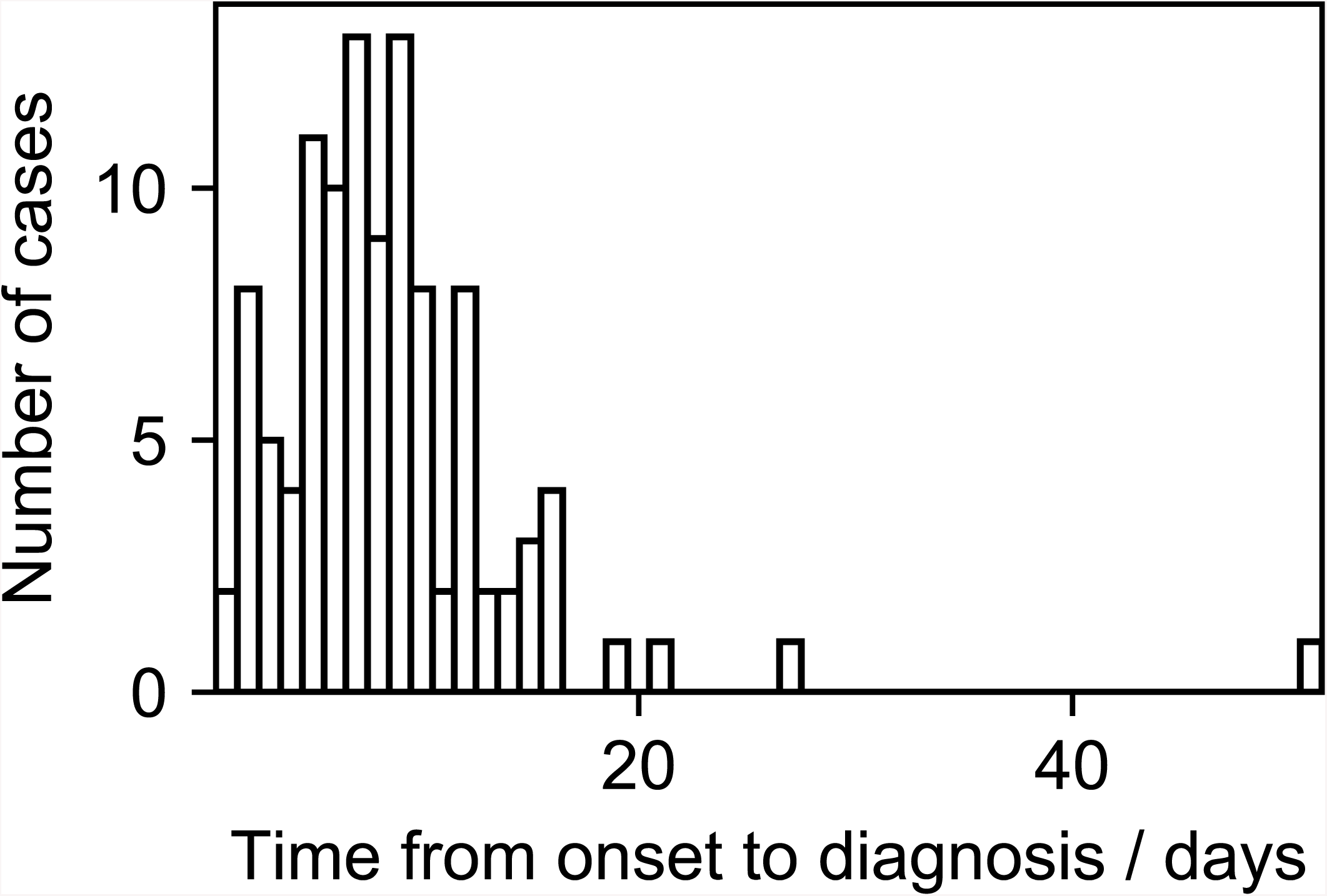
Distribution of the time from onset to diagnosis in dataset 1 and 2.

## Data Availability

All data referred to in the manuscript are attached as Supporting Table 1.

## Supporting information

**S1 Table. The cases of COVID-19 in Hokkaido, Japan**.

## Acknowledgments

This work was partially supported by JSPS KAKENHI Grant Number JP19K19429.

## Notes

### Competing Interest Statement

The authors have declared no competing interest.

